# Diagnostic Yield from Screening and Health Status Burden of Outpatients at Risk for Heart Failure

**DOI:** 10.1101/2025.02.25.25322868

**Authors:** Omar Cantu-Martinez, Andrew A. Girard, Weiwei Jin, Derek Rinderknecht, Thomas Cheek, John A. Spertus

**Author notes:** **Address for correspondence (Corresponding Author): Omar Cantu Martinez, MD**, Department of Cardiology, Saint Luke’s Mid America Heart Institute, 4401 Wornall Road, CV Research 9^th^ Floor Kansas City, MO 64111, Institutional. **Funding:** This study was funded by Ventric Health. **Disclosures:** Dr. Cantu Martinez receives funding from NHLBI award number T32HL110837-13 Dr. Girard receives funding from NHLBI award number T32HL110837. Dr. Jin discloses providing consultative services to and incentive compensation with Ventric Health. Dr. Rinderknecht discloses providing consultative services to and incentive compensation with Ventric Health. Dr. Cheek discloses providing consultative services to and incentive compensation with Ventric Health. Dr. Spertus discloses providing consultative services on patient-reported outcomes and evidence evaluation to Alnylam, AstraZeneca, Bayer, Janssen, Bristol Meyers Squibb, Terumo, Cytokinetics, BridgeBio, Ventric Health, and Imbria. He holds research grants from the National Institutes of Health, the Patient-Centered Outcomes Research Institute, the American College of Cardiology Foundation, Lexicon, Imbria, and Janssen. He owns the copyright to the Seattle Angina Questionnaire, Kansas City Cardiomyopathy Questionnaire, and Peripheral Artery Questionnaire and serves on the Board of Directors for Blue Cross Blue Shield of Kansas City.

## Abstract

**Background:** Heart failure (HF) is frequently underrecognized in primary care due to nonspecific symptoms and limited screening, resulting in many patients presenting with severely compromised health status (symptoms, functional ability, and quality of life) at the time of diagnosis.

**Objectives:** To evaluate the diagnostic yield of screening outpatients at risk for HF using a noninvasive assessment of left ventricular end-diastolic pressure (LVEDP) and to describe the health status of patients newly identified with elevated LVEDP.

**Methods:** A convenience sample of adults with diabetes mellitus (DM), chronic kidney disease (CKD), or suspected HF were screened at three primary care clinics using the Vivio System to identify patients with LVEDP >18 mmHg (positive screening). Among patients with a positive screening result, their health status was evaluated using the Kansas City Cardiomyopathy Questionnaire Overall Summary (KCCQ-OS) score.

**Results:** Among 2040 screened patients (mean age 74±8 years; 49.8% women; 64.6% with DM; and 34.9% with CKD) 38.5% had an elevated LVEDP. Older patients, women, and those with CKD were more likely to have an elevated LVEDP (p<0.01 for all). Of 653 KCCQ-OS scores collected (mean 85±20), 31.4% had a KCCQ-OS of 100 (asymptomatic), and 26.5% had a KCCQ-OS <80, consistent with NYHA class II-IV.

**Conclusion:** Nearly 40% of patients had a positive screening, and over two-thirds reported significant health status impairments. Combining the KCCQ with noninvasive LVEDP assessment can identify patients who may require further HF evaluation. Future studies can assess the impact of these strategies on patients’ subsequent health status and clinical events.

## INTRODUCTION

While the prevalence of Heart failure (HF) continues to rise,^1^ diagnosing it in primary care is challenging, particularly among elderly patients with multiple comorbidities ^2,3^ and nonspecific symptoms often attributed to aging or comorbidities ^4^. Earlier recognition could support initiating effective therapies to slow progression, reduce mortality, and improve patients’ health status.

Previous studies to improve HF diagnosis often rely upon tools not routinely available in primary care ^2,3^. The Vivio System was recently cleared by the US Food and Drug Administration (FDA) to noninvasively screen for elevated left ventricular end-diastolic pressure (LVEDP) ^5^. This report describes the real-world diagnostic yield of the Vivio System in outpatients and examines HF-specific health status at the time of elevated LVEDP detection to identify patients’ symptom burden at the time of diagnosis.

## METHODS

### Study Population and Design

Three primary care sites implemented the Vivio System (Ventric Health, Pasadena, CA) in routine clinical practice between August 13, 2024, and November 11, 2024. Adults with diabetes mellitus (DM), chronic kidney disease (CKD) stage ≥3, or a clinical suspicion for HF were encouraged to be screened. Patients with known HF or contraindications to the Vivio System (intravascular access, arterio-venous shunt/fistula, mastectomy, or lymph node dissection on the involved arm and any implantable electrical cardiac device) were excluded. Recorded comorbidities included hypertension, DM, CKD, chronic obstructive pulmonary disease, and arrhythmias. The 12-item Kansas City Cardiomyopathy Questionnaire (KCCQ) was recommended for patients identified as having an elevated LVEDP (positive screening) ^6^. As this was an analysis of de-identified data obtained from routine clinical care, it was reviewed by the Saint Luke’s Institutional Review Board and deemed to be non-human subjects research for which patient-level consent was not required.

### Vivio System

The 510k FDA-cleared Vivio System uses a modified pneumatic brachial blood pressure cuff that re-inflates to a supra-systolic blood pressure (+35 mmHg from systolic) to collect 40 seconds of brachial pulse waveform that is synchronized with a single-lead electrocardiogram to detect elevated LVEDP (>18 mmHg) via a classification model with 80% sensitivity and 83% specificity ^5^.

### Health Status Assessments

The KCCQ is a validated patient-reported measure that quantifies symptoms, function, and quality of life of HF patients. The Overall Summary (OS) score quantifies the full impact of HF on patients’ health status. The 12-item questionnaire (KCCQ-12) was used, which retains the psychometric properties of the original 23-item version ^6^. KCCQ-OS scores of 0–44, 45–59, 60– 79, and ≥80 roughly correspond to New York Heart Association (NYHA) functional classes IV, III, II, and I, respectively ^7^.

### Statistical Analysis

Continuous variables are reported as mean ± standard deviation and categorical variables as counts (%). T-tests and chi-squared tests were used to compare the characteristics of patients with and without elevated LVEDP. Ages >90 were imputed as 90 to comply with the Health Insurance Portability and Accountability Act. The distribution of KCCQ-OS scores (100, 80–99, 60–79, and <60) were described. Analyses were conducted by Ventric Health, with oversight by all authors, using Python 3.9.13 with pandas 1.4.3 and scipy 1.13.1.

## RESULTS

### Population Characteristics and Diagnostic Yield among Screened Patients

The overall study cohort included 2040 screened patients. The mean age was 74±8 years; 1015 (49.8%) were women, 1318 (64.6%) had a history of DM, and 711 (34.9%) had CKD. Among screened patients, 785 (38.5%) had an elevated LVEDP. Patients with an elevated LVEDP were older (74±8 versus 73±8, <0.01), more often women (62.2% versus 42.0%, p<0.001), and more likely to have CKD (38.3% versus 32.7%, p=0.01; Figure 1).

**Figure 1.**
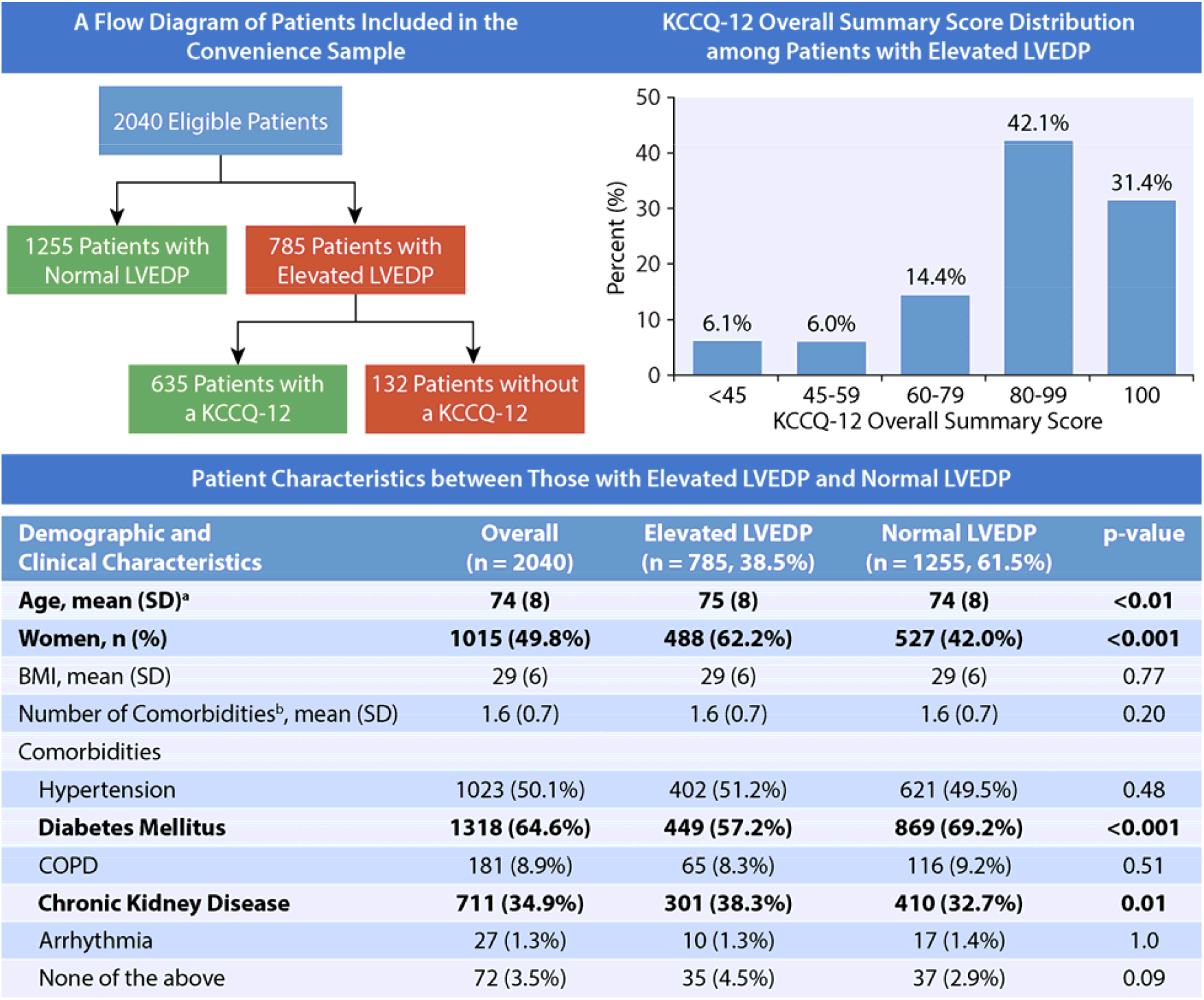
Central illustration. Cohort characteristics and health status of patients with elevated LVEDP. Abbreviations: KCCQ: Kansas City cardiomyopathy questionnaire, LVEDP: left ventricular end-diastolic pressure. *Caption: This figure summarizes the flow diagram of included patients, the health status distribution of patients with elevated left ventricular end-diastolic pressure measured by the Kansas City cardiomyopathy questionnaire, and patient characteristics of those patients with elevated left ventricular end-diastolic pressure (LVEDP) compared to those with normal LVEDP*. LVEDP: left ventricular end-diastolic pressure, KCCQ: Kansas City cardiomyopathy questionnaire, SD: standard deviation, BMI: body mass index, COPD: chronic obstructive pulmonary disease. ^a^ Patients with age ≥90 were imputed as 90 as the exact age was unavailable due to HIPPA privacy rules. ^b^ Only counting hypertension, diabetes mellitus, COPD, chronic kidney disease, and arrhythmia.

### Health Status of patients with elevated LVEDP at the time of diagnosis

Among the 785 patients with an elevated LVEDP, 653 (83.2%) completed the KCCQ-12. Almost a third, 205 (31.4%), had KCCQ-OS scores of 100, suggesting that they were asymptomatic (American Heart Association (AHA) stage B, pre-HF). Of those with AHA stage C HF, 275 (42.1%) had scores of 80-99, 94 (14.4%) had scores of 60-79, and 79 (12.1%) had scores <60, consistent with NYHA Class I, II, and III/IV, respectively.

## DISCUSSION

Despite advances in HF treatment, undiagnosed patients do not receive them. To overcome diagnostic delays in primary care, the FDA-cleared Vivio System was implemented as part of a screening protocol in patients with HF risk factors and no prior diagnosis. In this first-ever report, over one-third of screened patients had an elevated LVEDP, consistent with the diagnostic yield of previous efforts to screen high-risk elderly outpatients with comorbidities ^2,3^. Over two-thirds were symptomatic, with a quarter likely to be NYHA Class II-IV and at elevated risk for HF hospitalization and death ^8^. These data highlight a novel strategy to improve HF diagnoses in primary care. However, additional studies are needed to define the proportion confirmed to have HF and the impact of diagnosis on subsequent care and outcomes.

This study supports and extends the extant literature on screening high-risk patients for HF ^2,3^. Moreover, a higher proportion of women had elevated LVEDP, reinforcing previous research suggesting delayed HF diagnoses, lower treatment rates, and poorer health status compared with men ^9,10^. Future studies should also assess whether systematic HF screening can reduce sex-, race-, and socioeconomic-associated disparities in HF care and outcomes.

### Study Limitations

This study should be interpreted in the context of several potential limitations. It used a convenience sample from three primary care practices, limiting generalizability. The completeness of screening was not assessed and could inform future implementation strategies. Finally, HF was not independently confirmed despite elevated LVEDP and HF symptoms (KCCQ-OS), nor could these cross-sectional data assess treatment changes or outcomes after detecting elevated LVEDP.

## CONCLUSION

Considering the growing prevalence of HF and evolving treatments, there is a need for earlier diagnosis in primary care. In this initial experience with the Vivio System, nearly 40% of patients had a positive screening result, with over two-thirds having significant health status impairment. Combining the KCCQ with noninvasive LVEDP assessment could help identify patients who may benefit from further HF evaluation and treatment, potentially improving their health status and reducing clinical events.

## Data Availability

All data produced in the present study are available upon reasonable request to the authors

## Acknowledgments

None.

## Abbreviations

HF: heart failure
FDA: Food and Drug Administration
LVEDP: left ventricular end-diastolic pressure
DM: diabetes mellitus
CKD: chronic kidney disease
KCCQ: Kansas City Cardiomyopathy Questionnaire
OS: overall summary
NYHA: New York Heart Association
AHA: American Heart Association

